# Depression and incidence of inflammation-related physical health conditions: a cohort study in UK Biobank

**DOI:** 10.1101/2025.01.16.25320668

**Authors:** Shuvajit Saha, Regina Prigge, Caroline A Jackson, Bruce Guthrie, Kelly J Fleetwood

**Affiliations:** Usher Institute, University of Edinburgh, Edinburgh, UK; Advanced Care Research Centre, Usher Institute, University of Edinburgh, Edinburgh, UK

**Author notes:** Corresponding author: Shuvajit Saha.

**Keywords:** Depression, Inflammation, Physical health, Coronary Heart Disease, Peripheral Arterial Disease, Type 2 Diabetes, Inflammatory Bowel Disease, Inflammatory Arthritis, Parkinson’s Disease, UK Biobank

## Abstract

**Background:** Depression is associated with multiple physical health conditions, and inflammation is a mechanism commonly proposed to explain this association. We aimed to investigate the association between depression and the incidence of physical health conditions thought to have an inflammatory etiological component, including coronary heart disease, peripheral arterial disease, type 2 diabetes, inflammatory bowel disease, inflammatory arthritis and Parkinson’s Disease.

**Methods:** We conducted a cohort study using UK Biobank (UKB) data linked to primary care, hospital admission and death data. We ascertained depression at baseline using primary care and hospital records, and self-report at the UKB baseline assessment. We identified incident physical health conditions during follow-up using primary care, hospital admission and death data. We used Cox proportional hazards models to determine hazard ratios of each incident inflammation-related condition in those with versus without depression at baseline, serially adjusting for sociodemographic factors, lifestyle factors and baseline count of morbidities.

**Result:** We included 172,556 UKB participants who had continuous primary care records. Of these, 30,770 (17.8%) had a history of depression at baseline. After excluding participants with missing data, 168,641 (98%) were included in analysis. Median follow-up was 7.1 years (IQR: 6.3, 8.0). In the model adjusted for age and sex, depression was significantly associated with a higher hazard of all inflammation-related conditions. After additionally accounting for differences in country, ethnicity and deprivation, the association between depression and each condition generally attenuated but remained statistically significant, with effect estimates ranging from a 30% increased hazard of inflammatory bowel disease (HR 1.30, 95% CI 1.06 to 1.58) to a 53% increased hazard of Parkinson’s Disease (HR 1.53, 95% CI 1.25 to 1.87). After further adjusting for lifestyle factors and comorbidity count, the association persisted only for Parkinson’s Disease (HR 1.45, 95% CI 1.18 to 1.79).

**Conclusions:** Our study found that depression is consistently associated with multiple inflammation-related physical health conditions, although associations did not persist after adjustment for lifestyle factors and baseline physical condition count. Further research is needed to explore underlying mechanisms, including inflammatory biomarkers and modifiable lifestyle factors on the causal pathway.

## Background

Depression is a common mental health condition, affecting an estimated 300 million people worldwide [1]. The Global Burden of Disease Study 2019 reported an age-standardized prevalence of depressive disorders of 3440 per 100,000 people, with depressive disorders ranked 13^th^ for disease burden, worldwide [2]. As well as diminishing human capital and reducing quality of life [1], there is growing evidence that depression may be an independent risk factor for the development of some physical health conditions. Evidence from meta-analyses indicate that depression is associated with an 81% higher risk of developing coronary heart disease (CHD) [3], a 60% increased risk of developing type 2 diabetes (T2D) [4] and an increased risk of stroke [5], inflammatory bowel disease (IBD) [6] and Parkinson’s disease (PD) [7].

The link between depression and physical health conditions is likely to be multifactorial, but evidence suggests that chronic low-grade inflammation plays a crucial role in this complex association [8]. In a recent case-control study of UK Biobank (UKB) participants, individuals with depression versus without were found to have higher systemic inflammation levels [9]. Moreover, evidence indicates that depression may precede and augment pro-inflammatory cytokines, including interleukin-6 (IL-6) and C-reactive protein (CRP), potentially contributing to the development of cardiometabolic and other age-related diseases in healthy older adults [10]. Studies have also shown that anti-inflammatory agents can positively affect depressive symptoms [11]. Depression itself may therefore have an underlying inflammatory aetiological component and/or may lead to an altered inflammatory profile, each of which could help to explain a link with physical health conditions.

If the link between depression and physical health conditions is at least partly due to an inflammatory mechanism, then we would expect to observe an association between depression and the incidence of various inflammation-related conditions. However, the data supporting depression as a risk factor for some inflammation-related conditions, particularly peripheral arterial disease (PAD) and inflammatory arthritis (IA), is less robust, primarily due to the lack of large cohort studies. Moreover, to our knowledge, no study has examined associations between depression and incidence of multiple physical health conditions associated with inflammation within a single population. Therefore, we aimed to investigate the association between depression and the incidence of various conditions thought to have an inflammatory aetiological component among mid-aged participants within the UKB cohort.

## Methods

### Study design and participants

We conducted a cohort study using UKB data. The UKB is a cohort of half a million mid-aged adults with information on a wide range of health conditions [12]. Individuals aged 40-69 years and registered with a general practitioner in England, Scotland or Wales were invited. Participants attended baseline assessments between 2006 and 2010, which involved a touch-screen questionnaire, verbal interview, and physical measurements [12,13]. Participants provided written informed consent to follow-up through linkage to national datasets including primary care, hospital, cancer registry and death records. UKB has ethical approval from the NHS North West Research Ethics Committee (reference: 21/NW/0157).

The present study population comprised UKB participants with linked primary care data who had continued consent to participation. Methods are summarised here, and described in detail in a previous paper [14]. Linked primary care data was available from Scotland, Wales and practices in England that used either the SystmOne or Vision practice management systems. We included individuals with a continuous primary care record (no gaps of more than 90 days between practice registrations) from at least one year prior to their baseline assessment and extending to at least one day after the baseline assessment [15]. We excluded the small proportion of primary care records from the UKB extract of the Vision practice management system in England because this linked dataset is missing records from people who died before data extraction.

### Linked electronic health records

We ascertained the presence of conditions at baseline using information provided by the participants during their baseline assessment as well as from primary care, hospital, and cancer registry records. We identified conditions from primary care records using Read V2 and Clinical Terms V3 (CTV3) codes, from hospital records using ICD-10 codes and OPCS-4 procedure codes, and from death records using ICD-10 codes. All code lists are available in our project GitHub repository (https://github.com/rprigge-uoe/mltc-codelists). When people relocate within the UK, their primary care records are transferred between practices, and thus their records should capture their entire medical history. Hence, we used all primary care records up to and including the date of the participant’s baseline assessment to define conditions at baseline. Cancer registry and hospital records were available from different dates for England, Wales, and Scotland, with a minimum of eight years of records prior to the baseline assessments. To maintain consistency across secondary care data sources and the three countries, conditions at baseline were defined for each participant using cancer registry and hospital records spanning eight years up to and including their baseline assessment date.

We ascertained the occurrence of incident physical health conditions during follow-up from primary care, hospital and death records. Health record follow-up varied by country, with primary care and cancer registry records available up to at least 2016, while hospital and death records were complete to 2022. Participants were therefore followed up until the earliest of death, the end of continuous primary care records, or the end of cancer registry follow-up (Additional file 1: Table S1).

### Depression

A history of depression at baseline was identified if a participant had a prior diagnosis of depression in their linked primary care or hospital records, or if they self-reported a history of depression in response to the baseline assessment question, “Has a doctor ever told you that you have had any other serious medical conditions or disabilities?”

### Inflammation-related physical health conditions

We identified incident physical health conditions associated with inflammation (amongst people without each specific condition at baseline). Two clinicians [SS, BG] selected six conditions/groups of conditions associated with inflammation from a list of 80 long-term health conditions (appendix 2): coronary heart disease (CHD), peripheral arterial disease (PAD), type 2 diabetes (T2D), inflammatory bowel disease (IBD), Parkinson’s Disease (PD), and inflammatory arthritis and related conditions (IA). CHD was defined as including myocardial infarction, stable angina, unstable angina and CHD not otherwise specified. IA and related conditions comprised ankylosing spondylitis, juvenile arthritis, lupus erythematosus (local and systemic), polymyalgia rheumatica, post infective and reactive arthropathies, psoriatic arthropathy, rheumatoid arthritis, and systemic sclerosis (henceforth just called IA).

### Covariates

Age at baseline assessment and sex were determined from recruitment data and optionally updated by participants at the baseline assessment. Self-reported ethnicity was categorized into three groups: White, South Asian, and other ethnic minority groups. Country of residence (England, Wales, or Scotland) and area-based deprivation, assessed through the Townsend Deprivation Index [16], were derived from participants’ home addresses at baseline. The Townsend Deprivation Index was divided into deciles across the entire UKB cohort, ranging from 1 (least deprived) to 10 (most deprived).

Information on smoking, alcohol intake frequency, sleep disturbance, and physical activity was collected from the baseline assessment touchscreen questionnaire. Body mass index (BMI) was obtained from measurements taken during the baseline assessment. BMI was classified according to World Health Organization (WHO) guidelines, including the following categories: BMI <25 kg/m², BMI 25-29.9 kg/m², BMI 30-34.9 kg/m², and BMI ≥35 kg/m². We defined smoking status as current, previous and never smokers [17]. Participants were asked about their alcohol consumption frequency, with responses ranging from daily or almost daily to never. Sleep disturbance was determined from responses to the question ‘Do you have trouble falling asleep at night or do you wake up in the middle of the night?’ with responses of never/rarely, sometimes or usually. Additionally, low physical activity was included as a dichotomous variable, defined as engaging in no activity or light activity with a frequency of once per week or less [18]. For each participant, we counted the number of long-term health conditions (Additional file 1: Table S2) at baseline including 69 physical health conditions and 10 mental health conditions. Conditions were identified from the baseline assessment and from primary care, hospital and cancer registry records, as described above.

### Statistical analysis

For each of the six outcome conditions, we used Cox proportional hazard models to estimate the hazard ratios (HRs) with 95% confidence intervals (CIs) for time to incidence of the inflammation-related condition(s) by depression status at baseline. For each of the six outcome condition, individuals with that condition at baseline were excluded from the analysis. We obtained crude (unadjusted) estimates, with subsequent models adjusting for covariates as follows: model 1 adjusted for age at baseline and sex; model 2 additionally adjusted for other baseline socio-demographic factors, specifically country of residence, ethnicity, and Townsend Deprivation Index; and model 3 additionally adjusted for lifestyle factors (smoking, alcohol intake frequency, physical activity, sleep disturbance, and BMI) and baseline count of morbidities. Age and number of conditions at baseline were included in the models as continuous variables, each with a linear term and a quadratic term. Age and number of conditions at baseline were included in the models as continuous variables, scaled by subtracting their respective means. For each of these variables, we included the linear term, and we additionally included a quadratic term if it improved the fit of the model. The proportional hazards assumption was checked for all variables included in the study by using log cumulative hazard plots. The analysis was conducted using R version 4.3.2 [19]. Each covariate had less than 1% missing data (table 1) and 98% of participants had complete data. We therefore conducted a complete-case analysis rather than performing multiple imputation.

**Table 1:** Baseline characteristics for participants.

| Baseline characteristics | Depression<br>(N=30770) | No depression<br>(N=141786) | Total<br>(N=172556) |
| --- | --- | --- | --- |
| Age (years), mean (SD) | 56.3 (7.9) | 56.8 (8.0) | 56.7 (8.0) |
| Female (%) | 20592 (66.9) | 73431 (51.8) | 94023 (54.5) |
| Ethnicity (%) |  |  |  |
| White | 29620 (96.3) | 135086 (95.3) | 164706 (95.5) |
| South Asian | 363 (1.2) | 2440 (1.7) | 2803 (1.6) |
| Ethnic minority groups | 646 (2.1) | 3658 (2.6) | 4304 (2.5) |
| Missing | 141 (0.5) | 602 (0.4) | 743 (0.4) |
| Country (%) |  |  |  |
| England | 23709 (77.1) | 107911 (76.1) | 131620 (76.3) |
| Scotland | 3617 (11.8) | 18497 (13.0) | 22114 (12.8) |
| Wales | 3429 (11.1) | 15335 (10.8) | 18764 (10.9) |
| Missing | 15 (0.05) | 43 (0.03) | 58 (0.03) |
| Townsend deprivation index (%) |  |  |  |
| 1 (least deprived) | 2616 (8.5) | 14386 (10.1) | 17002 (9.9) |
| 2 | 2740 (8.9) | 15490 (10.9) | 18230 (10.6) |
| 3 | 2870 (9.3) | 15018 (10.6) | 17888 (10.4) |
| 4 | 2711 (8.8) | 14245 (10.0) | 16956 (9.8) |
| 5 | 3083 (10.0) | 15036 (10.6) | 18119 (10.5) |
| 6 | 3052 (9.9) | 14833 (10.5) | 17885 (10.4) |
| 7 | 3167 (10.3) | 14207 (10.0) | 17374 (10.1) |
| 8 | 3264 (10.6) | 13955 (9.8) | 17219 (10.0) |
| 9 | 3498 (11.4) | 13265 (9.4) | 16763 (9.7) |
| 10 (most deprived) | 3722 (12.1) | 11192 (7.9) | 14914 (8.6) |
| Missing | 47 (0.2) | 159 (0.1) | 206 (0.1) |
| Total number of conditions at baseline, median (IQR) | 3 (2.0, 5.0) | 2 (1.0, 3.0) | 2 (1.0, 3.0) |
| Inflammation-related conditions at baseline (%) |  |  |  |
| Coronary heart disease | 2680 (8.7) | 9486 (6.7) | 12166 (7.1) |
| Peripheral arterial disease | 379 (1.2) | 1220 (0.9) | 1599 (0.9) |
| Type 2 diabetes | 1766 (5.7) | 6180 (4.4) | 7946 (4.6) |
| Inflammatory bowel disease | 597 (1.9) | 2077 (1.5) | 2674 (1.5) |
| Parkinson's disease | 75 (0.24) | 240 (0.17) | 315 (0.18) |
| Inflammatory arthritis & other infl diseases* | 1337 (4.3) | 4351 (3.1) | 5688 (3.3) |
| Smoking (%) |  |  |  |
| Never | 15116 (49.1) | 79952 (56.4) | 95068 (55.1) |
| Previous | 10972 (35.7) | 48005 (33.9) | 58977 (34.2) |
| Current | 4524 (14.7) | 13128 (9.3) | 17652 (10.2) |
| Missing | 158 (0.5) | 701 (0.5) | 859 (0.5) |
| Alcohol intake (%) |  |  |  |
| Never | 3394 (11.0) | 10493 (7.4) | 13887 (8.0) |
| Special occasions only | 4566 (14.8) | 14777 (10.4) | 19343 (11.2) |
| 1-3 times a month | 3850 (12.5) | 15543 (11.0) | 19393 (11.2) |
| 1-2 times a week | 7629 (24.8) | 38156 (26.9) | 45785 (26.5) |
| 3 or 4 times a week | 5832 (19.0) | 34203 (24.1) | 40035 (23.2) |
| Daily or almost daily | 5411 (17.6) | 28329 (20.0) | 33740 (19.6) |
| Missing | 88 (0.3) | 285 (0.2) | 373 (0.2) |
| Low physical activity (%) |  |  |  |
| No | 25934 (84.3) | 126933 (89.5) | 152867 (88.6) |
| Yes | 4441 (14.4) | 13654 (9.6) | 18095 (10.5) |
| Missing | 395 (1.3) | 1199 (0.8) | 1594 (0.9) |
| Sleep disturbance (%) |  |  |  |
| Never/rarely | 4747 (15.4) | 36011 (25.4) | 40758 (23.6) |
| Sometimes | 13820 (44.9) | 68121 (48.0) | 81941 (47.5) |
| Usually | 12122 (39.4) | 37337 (26.3) | 49459 (28.7) |
| Missing | 81 (0.3) | 317 (0.2) | 398 (0.2) |
| **BMI (kg/m<sup>2</sup>) (%)** |  |  |  |
| <25 | 8873 (28.8) | 46121 (32.5) | 54994 (31.9) |
| 25 - 29.9 | 12434 (40.4) | 60998 (43.0) | 73432 (42.6) |
| 30 - 34.9 | 6143 (20.0) | 24737 (17.4) | 30880 (17.9) |
| >= 35 | 3121 (10.1) | 9132 (6.4) | 12253 (7.1) |
| Missing | 199 (0.6) | 798 (0.6) | 997 (0.6) |

Abbreviation: BMI, Body Mass Index; IQR, Interquartile Range
\*Includes ankylosing spondylitis, juvenile arthritis, lupus erythematosus (local and systemic), polymyalgia rheumatica, post infective and reactive arthropathies, psoriatic arthropathy, rheumatoid arthritis, and systemic sclerosis.

## Results

Among the 172,556 participants who met the inclusion criteria, 30,770 (17.8%) had a history of depression at baseline (table 1). After excluding participants with missing data, 168,641 (98%) were included in statistical models (figure 1). Median follow-up was 7.1 years (IQR: 6.3, 8.0). The mean age at baseline was 57 years. Most participants were white and from England, with no difference in depression prevalence by ethnicity. Participants with depression were more likely to be female and more commonly lived in areas with high deprivation. Current and previous smoking, low physical activity, sleep disturbance, and obesity were more common in people with versus without depression. Individuals with depression were more likely not to drink alcohol or to drink alcohol only occasionally compared to those without depression. At baseline, people with depression had a median of one more health condition compared to those without depression, and had higher baseline prevalence of CHD, PAD, T2D, IBD, PD and IA.

**Figure 1:**
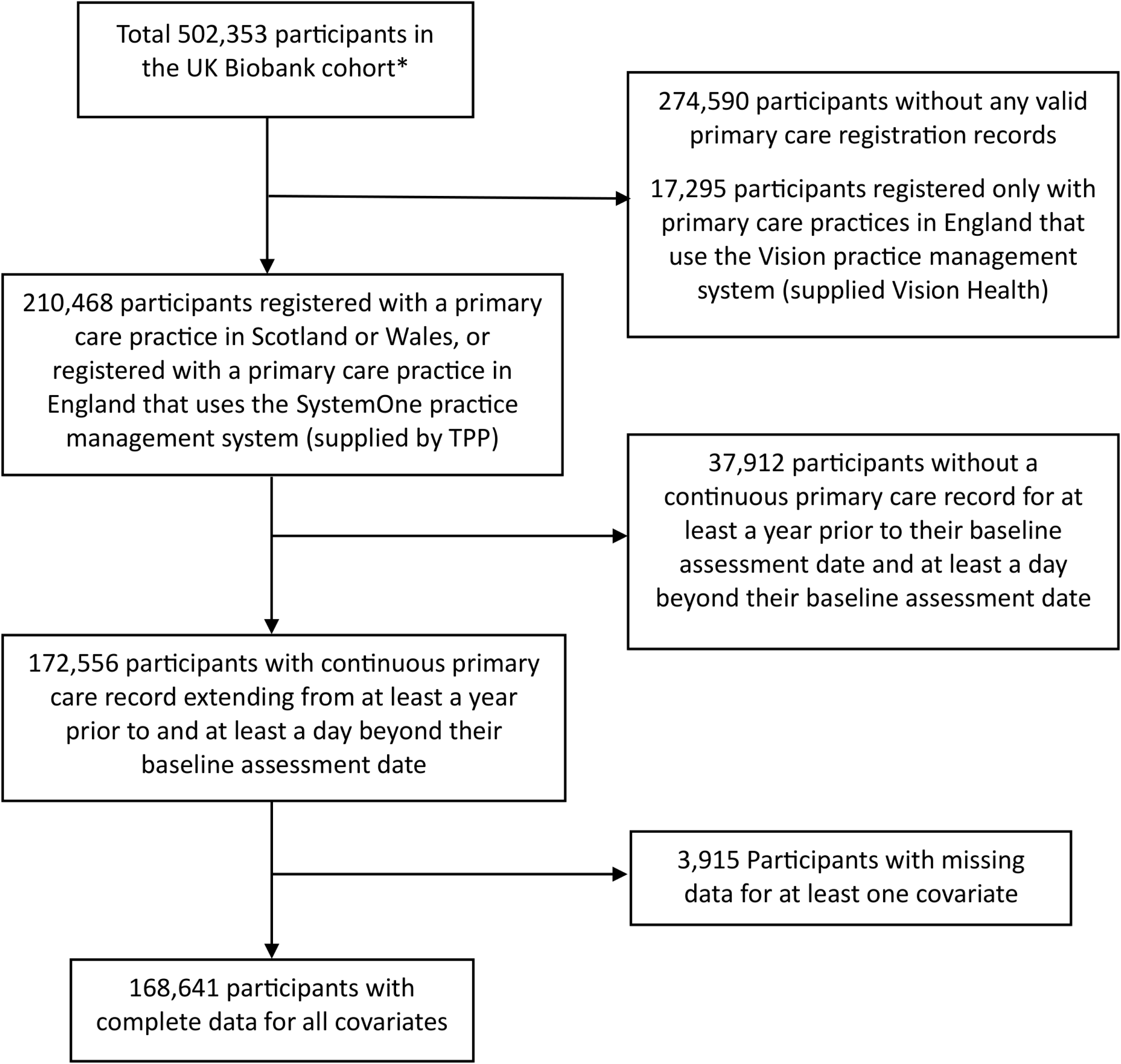
Flow diagram of UK Biobank sample selection. * Excluding participants who withdrew permission for their data to be included in research before 13 October 2023

Among individuals free of each inflammation-related condition at baseline, 4.6%, 3.7%, and 1.7% of participants developed CHD, T2D, and IA, respectively. However, fewer than 1% of participants developed PAD, IBD, and PD. Kaplan-Meier curves illustrate that participants with depression developed each condition at a faster rate than participants without depression (figure 2).

**Figure 2:**
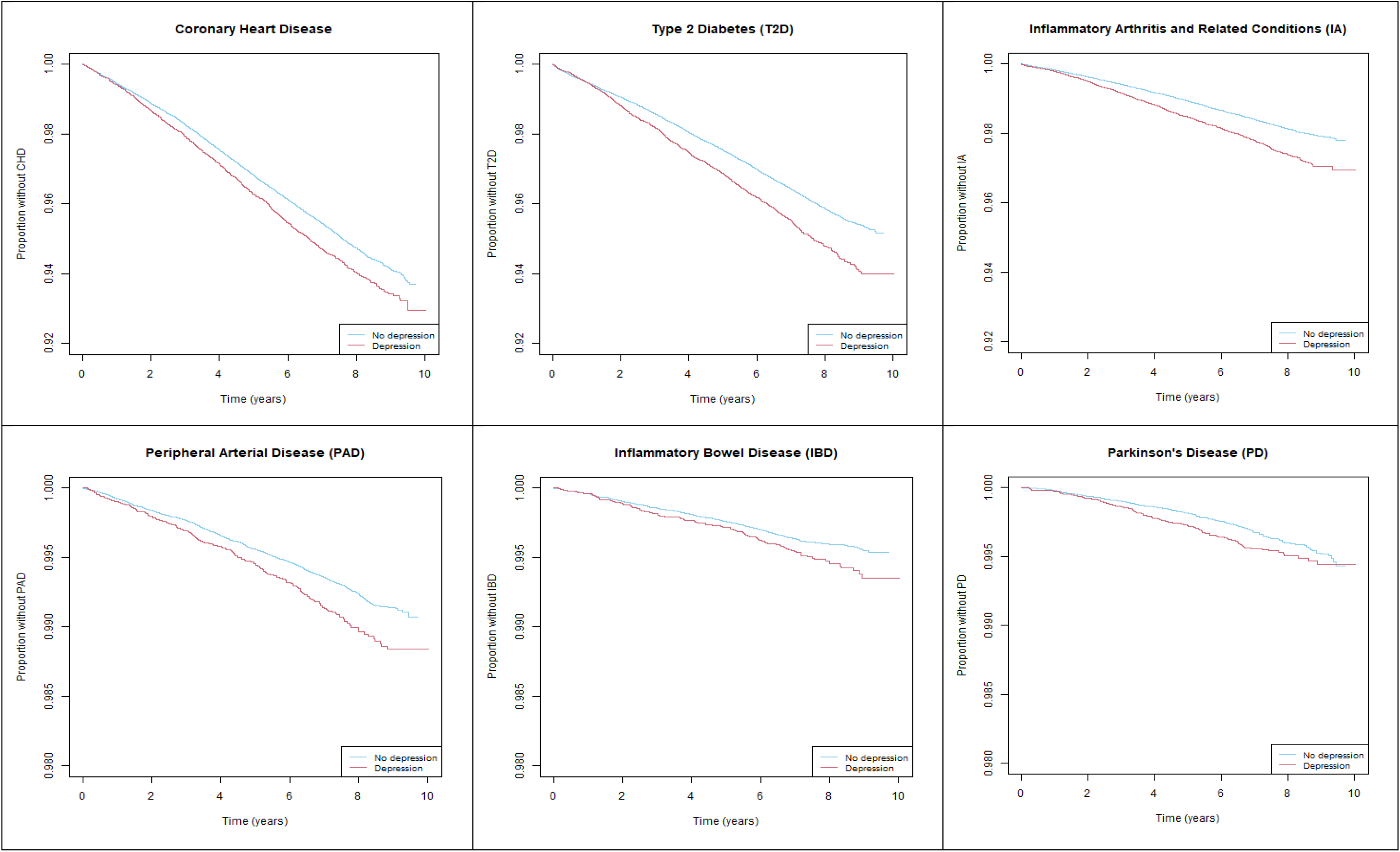
Proportion free of each inflammation-related condition over time, by depression status at baseline.

In the unadjusted model, a history of depression was associated with a statistically significantly higher hazard of all of the inflammation-related conditions (figure 3). After adjustment for age and sex, compared to people without a history of depression, people with a history of depression had a 36% increased hazard of CHD (HR 1.36, 95% CI 1.28–1.44), a 58% increased hazard of PAD (HR 1.58, 95% CI 1.37–1.83), a 42% increased hazard of T2D (HR 1.42, 95% CI 1.34–1.52), a 34% increased hazard of IBD (HR 1.34, 95% CI 1.10–1.63), a 52% increased hazard of PD (HR 1.52, 95% CI 1.25–1.86), and 37% increased hazard of IA (HR 1.37, 95% CI 1.25-1.50). Further accounting for country, ethnicity and deprivation made little difference to the estimated associations between depression and each condition. Following further adjustment for number of baseline comorbidities and lifestyle factors, there were no statistically significant associations between depression and five of the six conditions, the exception being PD (HR 1.45, 95% CI 1.18-1.79; figure 3).

**Figure 3:**
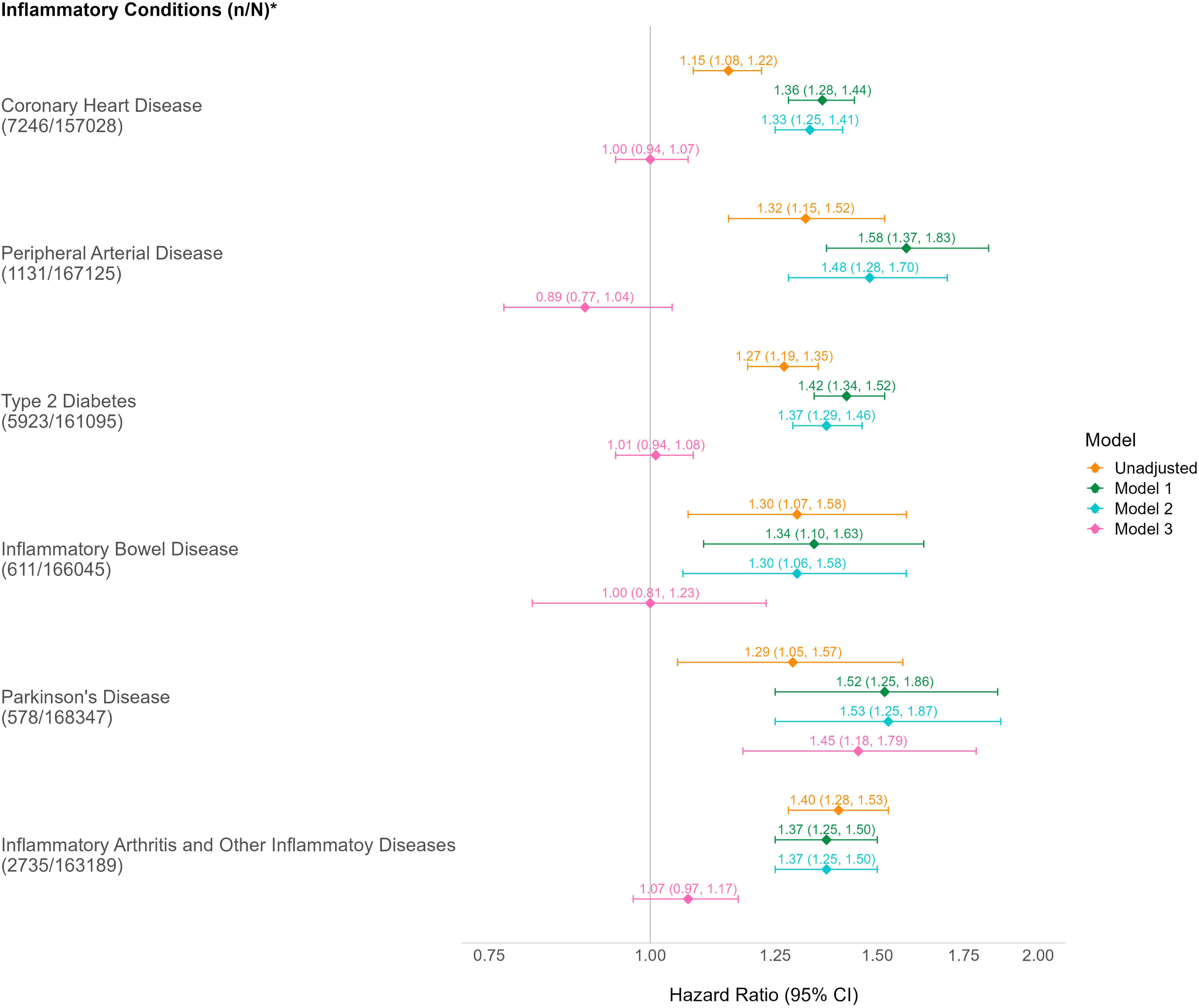
Unadjusted and adjusted hazard ratios (with 95% confidence intervals) for associations between depression and inflammation-related physical health conditions. * n= Number of participants developing the condition during follow-up; N= Number of participants without the condition at baseline Model 1: Adjusted for age and sex Model 2: Further adjusted for ethnicity, country, and Townsend deprivation index Model 3: Further adjusted for number of the conditions, smoking, alcohol intake, low physical activity, sleep disturbance, and BMI

## Discussion

We found that a history of depression at baseline was associated with an increased incidence of inflammation-related physical health conditions in middle and early old age. This association persisted after adjusting for age, sex and other sociodemographic factors. After additionally adjusting for baseline comorbidities and lifestyle factors the association persisted for PD only.

Previous research documents associations between depression and incidence of CHD [20–22], T2D [4,23–25], IA [25–28], IBD [6] and PD [7]. Most studies on IA have focused solely on rheumatoid arthritis, whereas our study included a broader set of IA conditions. Although few studies have investigated the association between depression and incident PAD, a recent UKB cohort study found that self-reported frequency of depressive symptoms prior to the baseline assessment in the previous two-week period were associated with a higher risk of developing PAD [29]. After adjusting for age, sex, and other sociodemographic factors, our findings are consistent with previous studies that made similar adjustments, reporting an increased risk of incident CHD [30–36], PAD [29,37], T2D [38–40], and PD [41,42] among people with depression. While the strength of the associations between depression and CHD [30,31,33–36], PAD [29,37], and T2D [38–40] was similar in some previous studies, others observed much higher HRs of 2.22 for CHD [32], and 3.24 [41] and 3.13 [42] for PD. Potential explanations for the observed differences in these studies include the use of varying methods to measure depression, such as the Centre for Epidemiologic Studies Depression Scale (CESD) [32], International Classification of Primary Care (ICPC) codes [42], and diagnoses made by psychiatrists [41].

A further potential reason for differences between studies is the use of varying adjustment sets. Our current study found that the association between depression and incident CHD, PAD, T2D, IBD, and IA did not persist after additional adjustments for number of baseline comorbidities, disrupted sleep, and lifestyle factors, aligning with some previous studies [25,33,34,37,40,43–49] but contrasting with others [4,20–22,24–28,50]. In line with previous studies [51,52], in our study the association between depression and PD persisted after adjustment for all covariates. However, reverse causation could partially explain these results; that is, patients with undiagnosed PD may develop depressive symptoms years before the motor symptoms become apparent [53].

In our study, the difference in effect estimates following conservative versus wider adjustment for covariates might lead to the conclusion that the association between depression and inflammation-related physical health conditions is simply due to confounding rather than depression being an independent risk factor. However, the final model adjusts for factors that may play a mediating rather than a confounding role. For example, depression might lead to low physical activity, high BMI and disturbed sleep, each of which are thought to promote chronic low-grade systemic inflammation [54–58], which in turn might mechanistically increase the risk of various chronic diseases, including those examined in this study. While our study is not inconsistent with the hypothesis that inflammation is the mechanism linking depression and many physical health conditions, it does not preclude the possibility that depression and inflammation-related physical conditions may share a common mechanism. For example, a dysregulated hypothalamic-pituitary-adrenal axis may separately lead to both depression and cardiometabolic disease, contributing to the association between depression and cardiometabolic diseases [59,60]. Alternatively, behavioural pathways, including smoking, may be additional or alternative mechanisms accounting for associations between depression and CHD [59] and PAD [61], whilst physical inactivity may partly explain links with T2D and CHD [59]. There is also some evidence [62] suggesting shared genetic variations between depression and a range of physical health conditions including IA and PD.

Our study has several strengths. The large study population, long follow-up period and breadth of data collected in the UKB allowed us to examine the links between depression and multiple inflammation-related physical health conditions within a single cohort. Our study further benefits from the use linked primary care data, as well as hospital records, to ascertain incident physical health conditions. Many of these outcomes, particularly T2D and IBD, are under-ascertained when relying solely on hospital admission data.

Our study has some limitations. Firstly, there are some missing values in the socio-demographic characteristics and lifestyle-related variables, but the proportion of missing values for these variables was less than 1%, and only about 2% of participants have any missing data. Therefore, a complete case analysis is unlikely to have introduced significant bias. Secondly, in our current study, we did not account for CRP, a well-established marker of chronic inflammation, as repeated measures of CRP data were unavailable. However, CRP levels can fluctuate markedly due to acute infection and other transient factors, meaning that single measurements of CRP may not reliably measure chronic inflammation. Finally, the UKB cohort primarily consists of individuals with white ethnicity and is subject to the healthy cohort effect [63], which may limit the generalizability of the findings to other populations. Despite this, the UKB is a large cohort with 500,000 participants, and we included approximately 170,000 participants in the current study, ensuring sufficient heterogeneity to detect associations between baseline characteristics and health outcomes [12]. Finally, whilst we adjusted for various demographic characteristics and lifestyle factors, as with any observational study there may be residual confounding through misclassification of included confounders and unknown confounders.

In the current study, the association between depression and incident inflammation-related conditions did not persist after fully adjusting for age, sex, other sociodemographic factors, baseline comorbidities, and lifestyle factors. However, some of these factors likely play a mediating rather than a confounding role, detailed exploration of which was beyond the scope of the current study. These findings underscore the need for lifestyle interventions among individuals with depression to mitigate risk factors for cardiometabolic diseases and other physical health conditions. Health practitioners should prioritize screening for risk factors of physical illnesses in people with depression, with particular attention to conditions with potential inflammatory causes. A more integrated approach to patient care combining mental health evaluations with proactive management of physical health risks could improve outcomes and promote holistic well-being. Furthermore, future studies should adopt methodologies such as mediation analysis to investigate and untangle the underlying mechanisms linking depression and inflammation-related physical health conditions, including analysis of inflammatory biomarkers. Exploring genetic and environmental interactions could also offer a deeper understanding of individual susceptibility to these conditions, paving the way for precision medicine approaches in mental health care.

## Conclusions

Our study found that depression is consistently associated with multiple inflammation-related physical health conditions, although associations did not persist after adjustment for lifestyle factors and baseline physical condition count, except for PD. Further research is needed to better understand the underlying mechanisms, including formal mediation analyses with a specific focus on inflammatory biomarkers and the contribution of modifiable lifestyle factors that may lie on the causal pathway.

## Data Availability

This study was conducted using data from the UK Biobank. Researchers can apply to access the UK Biobank data for health research in the public interest. The code lists used in this study are available from https://github.com/rprigge-uoe/mltc-codelists.

## List of abbreviations

BMI: Body Mass Index
CHD: Coronary Heart Disease
CI: Confidence Interval
CRP: C-Reactive Protein
CTV3: Clinical Terms V3
HRs: Hazard Ratios
IA: Inflammatory Arthritis and Related Conditions
IBD: Inflammatory Bowel Disease
IL-6: Interleukin-6
PAD: Peripheral Arterial Disease
PD: Parkinson’s Disease
T2D: Type 2 Diabetes
UKB: UK Biobank
WHO: World Health Organization

## Declarations

## Ethics approval and consent to participate

UKB has ethical approval from the NHS North West Research Ethics Committee (reference:21/NW/0157). All participants provided written and informed consent for data collection, analysis, and record linkage.

## Consent for publication

Not Applicable

## Competing interests

The authors declare that they have no competing interests.

## Funding

This study was funded by Medical Research Council/National Institute for Health Research (MC/S028013).

## Authors’ contributions

BG was responsible for the conception of the study and all authors contributed to the study design. RP and KF derived the analysis dataset. SS performed the analyses, and all authors contributed to interpretation of the findings. SS wrote the first draft of the manuscript, and all authors commented on manuscript drafts. The authors read and approved the final manuscript.

## Acknowledgements

The study was conducted using the UK Biobank Resource under application number 57213. The authors would like to thank the UK Biobank participants and the UK Biobank staff for their contributions to this study.

## Notes

### Competing Interest Statement

The authors have declared no competing interest.

### Author Declarations

UK Biobank has ethical approval from the NHS North West Research Ethics Committee (reference: 21/NW/0157).

## References

1. Patel V, Chisholm D, Parikh R, Charlson FJ, Degenhardt L, Dua T, et al. Addressing the burden of mental, neurological, and substance use disorders: key messages from Disease Control Priorities, 3rd edition. Lancet. 2016;387:1672–85.

2. GBD 2019 Mental Disorders Collaborators. Global, regional, and national burden of 12 mental disorders in 204 countries and territories, 1990-2019: a systematic analysis for the Global Burden of Disease Study 2019. Lancet Psychiatry. 2022;9:137–50.

3. Nicholson A, Kuper H, Hemingway H. Depression as an aetiologic and prognostic factor in coronary heart disease: a meta-analysis of 6362 events among 146 538 participants in 54 observational studies. Eur Heart J. 2006;27:2763–74.

4. Mezuk B, Eaton WW, Albrecht S, Golden SH. Depression and type 2 diabetes over the lifespan: a meta-analysis. Diabetes Care. 2008;31:2383–90.

5. Pan A, Sun Q, Okereke OI, Rexrode KM, Hu FB. Depression and risk of stroke morbidity and mortality: a meta-analysis and systematic review. JAMA. 2011;306:1241–9.

6. Piovani D, Armuzzi A, Bonovas S. Association of depression with incident inflammatory bowel diseases: a systematic review and meta-analysis. Inflamm Bowel Dis. 2024;30:573–84.

7. Wang S, Mao S, Xiang D, Fang C. Association between depression and the subsequent risk of Parkinson’s disease: a meta-analysis. Prog Neuro-Psychopharmacology Biol Psychiatry. 2018;86:186–92.

8. Berk M, Köhler-Forsberg O, Turner M, Penninx BWJH, Wrobel A, Firth J, et al. Comorbidity between major depressive disorder and physical diseases: a comprehensive review of epidemiology, mechanisms and management. World Psychiatry. 2023;22:366–87.

9. Pitharouli MC, Hagenaars SP, Glanville KP, Coleman JRI, Hotopf M, Lewis CM, et al. Elevated C-Reactive Protein in patients with depression, independent of genetic, health, and psychosocial factors: results from the UK Biobank. Am J Psychiatry. 2021;178:522–9.

10. Stewart JC, Rand KL, Muldoon MF, Kamarck TW. A prospective evaluation of the directionality of the depression-inflammation relationship. Brain Behav Immun. 2009;23:936–44.

11. Köhler O, Benros ME, Nordentoft M, Farkouh ME, Iyengar RL, Mors O, et al. Effect of anti-inflammatory treatment on depression, depressive symptoms, and adverse effects: a systematic review and meta-analysis of randomized clinical Trials. JAMA Psychiatry. 2014;71:1381–91.

12. Sudlow C, Gallacher J, Allen N, Beral V, Burton P, Danesh J, et al. UK Biobank: an open access resource for identifying the causes of a wide range of complex diseases of middle and old age. PLOS Med. 2015;12:e1001779.

13. UK Biobank. UK Biobank: Protocol for a large-scale prospective epidemiological resource. UKBB-PROT-09-06 (Main Phase). 2007;06:1–112.

14. Prigge, Regina and Fleetwood, Kelly J. and Jackson, Caroline A. and Mercer, Stewart and Kelly, Paul AT and Sudlow, Cathie and Norrie, John D. and Morales, Daniel R. and Smith, Daniel J. and Guthrie B. Robustly measuring multiple long-term health conditions using disparate linked datasets in UK Biobank. SSRN [Internet]. Available from: https://ssrn.com/abstract=4863974

15. Lewis JD, Bilker WB, Weinstein RB, Strom BL. The relationship between time since registration and measured incidence rates in the General Practice Research Database. Pharmacoepidemiol Drug Saf. 2005;14:443–51.

16. Townsend P. Deprivation. J Soc Policy. 1987;16:125–146.

17. UK Biobank: Data-Field 20116 [Internet]. [cited 2024 Nov 30]. Available from: https://biobank.ndph.ox.ac.uk/showcase/field.cgi?id=20116

18. Hanlon P, Nicholl BI, Jani BD, Lee D, McQueenie R, Mair FS. Frailty and pre-frailty in middle-aged and older adults and its association with multimorbidity and mortality: a prospective analysis of 493D737 UK Biobank participants. Lancet Public Heal. 2018;3:e323–32.

19. R Core Team. R: a language and environment for statistical computing. Vienna, Austria: R Foundation for Statistical Computing; 2023. https://www.R-project.org/.

20. Cao H, Zhao H, Shen L. Depression increased risk of coronary heart disease: a meta-analysis of prospective cohort studies. Front Cardiovasc Med. 2022;9:913888.

21. Gan Y, Gong Y, Tong X, Sun H, Cong Y, Dong X, et al. Depression and the risk of coronary heart disease: a meta-analysis of prospective cohort studies. BMC Psychiatry. 2014;14:371.

22. Wu Q, Kling JM. Depression and the risk of myocardial infarction and coronary death: a meta-analysis of prospective cohort studies. Medicine (Baltimore). 2016;95:e2815.

23. Birk JL, Kronish IM, Moise N, Falzon L, Yoon S, Davidson KW. Depression and multimorbidity: considering temporal characteristics of the associations between depression and multiple chronic diseases. Heal Psychol Off J Div Heal Psychol Am Psychol Assoc. 2019;38:802–11.

24. Rotella F, Mannucci E. Depression as a risk factor for diabetes: a meta-analysis of longitudinal studies. J Clin Psychiatry. 2013;74:31–7.

25. Frank P, Batty GD, Pentti J, Jokela M, Poole L, Ervasti J, et al. Association between depression and physical conditions requiring hospitalization. JAMA Psychiatry. 2023;80:690–9.

26. Lu M-C, Guo H-R, Lin M-C, Livneh H, Lai N-S, Tsai T-Y. Bidirectional associations between rheumatoid arthritis and depression: a nationwide longitudinal study. Sci Rep. 2016;6:20647.

27. Vallerand IA, Lewinson RT, Frolkis AD, Lowerison MW, Kaplan GG, Swain MG, et al. Depression as a risk factor for the development of rheumatoid arthritis: a population-based cohort study. RMD Open. 2018;4.

28. Lewinson RT, Vallerand IA, Lowerison MW, Parsons LM, Frolkis AD, Kaplan GG, et al. Depression is associated with an increased risk of psoriatic arthritis among patients with psoriasis: a population-based study. J Invest Dermatol. 2017;137:828–35.

29. Lee SN, Yun J-S, Ko S-H, Ahn Y-B, Yoo K-D, Her S-H, et al. Impacts of gender and lifestyle on the association between depressive symptoms and cardiovascular disease risk in the UK Biobank. Sci Rep. 2023;13:10758.

30. Hawkins MAW, Callahan CM, Stump TE, Stewart JC. Depressive symptom clusters as predictors of incident coronary artery disease: a 15-year prospective study. Psychosom Med. 2014;76:38–43.

31. Brunner EJ, Shipley MJ, Britton AR, Stansfeld SA, Heuschmann PU, Rudd AG, et al. Depressive disorder, coronary heart disease, and stroke: dose-response and reverse causation effects in the Whitehall II cohort study. Eur J Prev Cardiol. 2014;21:340–6.

32. Péquignot R, Tzourio C, Péres K, Ancellin M-L, Perier M-C, Ducimetière P, et al. Depressive symptoms, antidepressants and disability and future coronary heart disease and stroke events in older adults: the Three City Study. Eur J Epidemiol. 2013;28:249–56.

33. O’Brien EC, Greiner MA, Sims M, Hardy NC, Wang W, Shahar E, et al. Depressive Symptoms and risk of cardiovascular events in blacks: findings from the Jackson heart study. Circ Cardiovasc Qual Outcomes. 2015;8:552–9.

34. Sims M, Redmond N, Khodneva Y, Durant RW, Halanych J, Safford MM. Depressive symptoms are associated with incident coronary heart disease or revascularization among blacks but not among whites in the reasons for geographical and racial differences in stroke study. Ann Epidemiol. 2015;25:426–32.

35. Péquignot R, Dufouil C, Prugger C, Pérès K, Artero S, Tzourio C, et al. High Level of depressive symptoms at repeated study visits and risk of coronary heart disease and stroke over 10 Years in older adults: The Three-City Study. J Am Geriatr Soc. 2016;64:118–25.

36. Nabi H, Kivimäki M, Suominen S, Koskenvuo M, Singh-Manoux A, Vahtera J. Does depression predict coronary heart disease and cerebrovascular disease equally well? The health and social support prospective cohort study. Int J Epidemiol. 2010;39:1016–24.

37. Grenon SM, Hiramoto J, Smolderen KG, Vittinghoff E, Whooley MA, Cohen BE. Association between depression and peripheral artery disease: insights from the heart and soul study. J Am Heart Assoc. 2012;1:e002667.

38. Campayo A, de Jonge P, Roy JF, Saz P, de la Cámara C, Quintanilla MA, et al. Depressive disorder and incident diabetes mellitus: the effect of characteristics of depression. Am J Psychiatry. 2010;167:580–8.

39. Carnethon MR, Biggs ML, Barzilay JI, Smith NL, Vaccarino V, Bertoni AG, et al. Longitudinal association between depressive symptoms and incident type 2 diabetes mellitus in older adults: the cardiovascular health study. Arch Intern Med. 2007;167:802–7.

40. van den Akker M, Schuurman A, Metsemakers J, Buntinx F. Is depression related to subsequent diabetes mellitus? Acta Psychiatr Scand. 2004;110:178–83.

41. Shen C-C, Tsai S-J, Perng C-L, Kuo BI-T, Yang AC. Risk of Parkinson disease after depression: a nationwide population-based study. Neurology. 2013;81:1538–44.

42. Schuurman AG, van den Akker M, Ensinck KTJL, Metsemakers JFM, Knottnerus JA, Leentjens AFG, et al. Increased risk of Parkinson’s disease after depression: a retrospective cohort study. Neurology. 2002;58:1501–4.

43. Golden SH, Lazo M, Carnethon M, Bertoni AG, Schreiner PJ, Diez Roux A V, et al. Examining a bidirectional association between depressive symptoms and diabetes. JAMA. 2008;299:2751–9.

44. Ananthakrishnan AN, Khalili H, Pan A, Higuchi LM, de Silva P, Richter JM, et al. Association between depressive symptoms and incidence of Crohn’s disease and ulcerative colitis: results from the Nurses’ Health sStudy. Clin Gastroenterol Hepatol Off Clin Pract J Am Gastroenterol Assoc. 2013;11:57–62.

45. Rahman I, Humphreys K, Bennet AM, Ingelsson E, Pedersen NL, Magnusson PKE. Clinical depression, antidepressant use and risk of future cardiovascular disease. Eur J Epidemiol. 2013;28:589–95.

46. Majed B, Arveiler D, Bingham A, Ferrieres J, Ruidavets J-B, Montaye M, et al. Depressive symptoms, a time-dependent risk factor for coronary heart disease and stroke in middle-aged men: the PRIME Study. Stroke. 2012;43:1761–7.

47. Janszky I, Ahnve S, Lundberg I, Hemmingsson T. Early-onset depression, anxiety, and risk of subsequent coronary heart disease: 37-year follow-up of 49,321 young Swedish men. J Am Coll Cardiol. 2010;56:31–7.

48. Wulsin LR, Evans JC, Vasan RS, Murabito JM, Kelly-Hayes M, Benjamin EJ. Depressive symptoms, coronary heart disease, and overall mortality in the Framingham Heart Study. Psychosom Med. 2005;67:697–702.

49. Saydah SH, Brancati FL, Golden SH, Fradkin J, Harris MI. Depressive symptoms and the risk of type 2 diabetes mellitus in a US sample. Diabetes Metab Res Rev. 2003;19:202–8.

50. Frolkis AD, Vallerand IA, Shaheen A-A, Lowerison MW, Swain MG, Barnabe C, et al. Depression increases the risk of inflammatory bowel disease, which may be mitigated by the use of antidepressants in the treatment of depression. Gut. 2019;68:1606–12.

51. Ishihara-Paul L, Wainwright NWJ, Khaw K-T, Luben RN, Welch AA, Day NE, et al. Prospective association between emotional health and clinical evidence of Parkinson’s disease. Eur J Neurol. 2008;15:1148–54.

52. Fang F, Xu Q, Park Y, Huang X, Hollenbeck A, Blair A, et al. Depression and the subsequent risk of Parkinson’s disease in the NIH-AARP Diet and Health Study. Mov Disord. 2010;25:1157–62.

53. Weintraub D, Caspell-Garcia C, Simuni T, Cho HR, Coffey CS, Aarsland D, et al. Neuropsychiatric symptoms and cognitive abilities over the initial quinquennium of Parkinson disease. Ann Clin Transl Neurol. 2020;7:449–61.

54. Nimmo MA, Leggate M, Viana JL, King JA. The effect of physical activity on mediators of inflammation. Diabetes, Obes Metab. 2013;15:51–60.

55. Geffken DF, Cushman M, Burke GL, Polak JF, Sakkinen PA, Tracy RP. Association between physical activity and markers of inflammation in a healthy elderly population. Am J Epidemiol. 2001;153:242–50.

56. de Heredia FP, Gómez-Martínez S, Marcos A. Obesity, inflammation and the immune system. Proc Nutr Soc. 2012/03/20. 2012;71:332–8.

57. Ellulu MS, Patimah I, Khaza’ai H, Rahmat A, Abed Y. Obesity and inflammation: the linking mechanism and the complications. Arch Med Sci. 2017;13:851–63.

58. Irwin MR, Olmstead R, Carroll JE. Sleep Disturbance, sleep duration, and inflammation: A systematic review and meta-analysis of cohort studies and experimental sleep deprivation. Biol Psychiatry. 2016;80:40–52.

59. Gold SM, Köhler-Forsberg O, Moss-Morris R, Mehnert A, Miranda JJ, Bullinger M, et al. Comorbid depression in medical diseases. Nat Rev Dis Prim. 2020;6:69.

60. Milaneschi Y, Lamers F, Berk M, Penninx BWJH. Depression heterogeneity and its biological underpinnings: toward immunometabolic depression. Biol Psychiatry. 2020;88:369–80.

61. Wang W, Zhao T, Geng K, Yuan G, Chen Y, Xu Y. Smoking and the pathophysiology of peripheral artery disease. Front Cardiovasc Med. 2021;8:704106.

62. Mulugeta A, Zhou A, King C, Hyppönen E. Association between major depressive disorder and multiple disease outcomes: a phenome-wide Mendelian randomisation study in the UK Biobank. Mol Psychiatry. 2020;25:1469–76.

63. Fry A, Littlejohns TJ, Sudlow C, Doherty N, Adamska L, Sprosen T, et al. Comparison of sociodemographic and health-related characteristics of UK Biobank participants with those of the general population. Am J Epidemiol. 2017;186:1026–34.

