## Additional File 1 for "Depression and incidence of inflammation-related physical health conditions: a cohort study in UK Biobank"

Table S1: Availability periods of the electronic health records

| **Data source** | **Region** | **Data complete** |
| --- | --- | --- |
| Hospital admission | Scotland | Earliest record: Jan 1981  Complete to: 31 Aug 2022 |
|  | Wales | Earliest record: Jan 1998*  Complete to: 31 May 2022 |
|  | England | Earliest record: Apr 1997  Complete to: 31 Oct 2022 |
| Cancer registry | Scotland | Earliest record: 1957  Complete to: 30 Nov 2021 |
|  | Wales | Earliest record: 1971  Complete to: 31 Dec 2016 |
|  | England | Earliest record: 1971  Complete to: 31 Dec 2020 |
| Primary care | Scotland | Earliest record^1^: Aug 1937  Complete to: 31 Mar 2017 |
|  | Wales | Earliest record^#^: May 1940  Compete to: 31 Aug 2017 |
|  | England (TPP) | Earliest record^1^: Dec 1937  Complete to: 31 May 2016 |

* At the time of publication, the UK Biobank website (https://biobank.ndph.ox.ac.uk/showcase/exinfo.cgi?src=Data_providers_and_dates) lists the start date for Wales as 1991, but data records are not consistently available until January 1, 1998

\# Records prior to each participant’s date of birth were excluded.

Table S2: Long-term health conditions measured at baseline

| **Condition type** | **Condition** | **Condition includes** |
| --- | --- | --- |
| Physical | Benign neoplasm of brain and other CNS |  |
|  | Haematological malignancies | Hodgkin lymphoma |
|  |  | Leukaemia |
|  |  | Monoclonal gammopathy of undetermined significance (MGUS) |
|  |  | Multiple myeloma and malignant plasma cell neoplasms |
|  |  | Myelodysplastic syndromes |
|  |  | Non-Hodgkin lymphoma |
|  |  | Polycythaemia vera |
|  |  | Haematological malignancy - other |
|  | Non-melanoma skin malignancies |  |
|  | Solid organ malignancies | Primary malignancy - biliary tract |
|  |  | Primary malignancy - bladder |
|  |  | Primary malignancy - bone and articular cartilage |
|  |  | Primary malignancy - brain, other central nervous system and intracranial |
|  |  | Primary malignancy - breast |
|  |  | Primary malignancy - cervical |
|  |  | Primary malignancy - colorectal and anus |
|  |  | Primary malignancy - kidney and ureter |
|  |  | Primary malignancy - liver |
|  |  | Primary malignancy - lung and trachea |
|  |  | Primary malignancy - malignant melanoma |
|  |  | Primary malignancy - mesothelioma |
|  |  | Primary malignancy - multiple independent sites |
|  |  | Primary malignancy - oesophageal |
|  |  | Primary malignancy - oro-pharyngeal |
|  |  | Primary malignancy - ovarian |
|  |  | Primary malignancy - pancreatic |
|  |  | Primary malignancy - prostate |
|  |  | Primary malignancy - stomach |
|  |  | Primary malignancy - testicular |
|  |  | Primary malignancy - thyroid |
|  |  | Primary malignancy - uterine |
|  |  | Primary malignancy - other organs |
|  |  | Secondary malignancy - adrenal gland |
|  |  | Secondary malignancy - bone |
|  |  | Secondary malignancy - bowel |
|  |  | Secondary malignancy - brain, other central nervous system and intracranial |
|  |  | Secondary malignancy - liver and intrahepatic bile duct |
|  |  | Secondary malignancy - lung |
|  |  | Secondary malignancy - lymph nodes |
|  |  | Secondary malignancy - pleura |
|  |  | Secondary malignancy - retroperitoneum and peritoneum |
|  |  | Secondary malignancy - other organs |
|  | Cardiomyopathy | Dilated cardiomyopathy |
|  |  | Hypertrophic cardiomyopathy |
|  |  | Other cardiomyopathy |
|  | Conduction disorders and other arrhythmias | Atrioventricular block, complete |
|  |  | Sick sinus syndrome |
|  |  | Supraventricular tachycardia |
|  | Coronary heart disease | Coronary heart disease not otherwise specified |
|  |  | Myocardial infarction |
|  |  | Stable angina |
|  |  | Unstable angina |
|  | Heart valve disorders | Multiple valve disorder |
|  |  | Nonrheumatic aortic valve disorders |
|  |  | Nonrheumatic mitral valve disorders |
|  |  | Rheumatic valve disorder |
| Physical | Stroke | Intracerebral haemorrhage |
|  |  | Ischaemic stroke |
|  |  | Stroke not otherwise specified |
|  | Atrial fibrillation |  |
|  | Heart failure |  |
|  | Hypertension |  |
|  | Peripheral arterial disease |  |
|  | Primary pulmonary hypertension |  |
|  | Transient ischaemic attack |  |
|  | Chronic liver disease | Alcoholic liver disease |
|  |  | Autoimmune liver disease |
|  |  | Hepatic failure |
|  |  | Liver fibrosis, sclerosis and cirrhosis |
|  |  | Portal hypertension |
|  |  | Chronic viral hepatitis |
|  | Gastro-oesophageal reflux, gastritis and similar [abbreviated to gastro-oesophageal reflux disease (and similar) in the main text] | Barrett's oesophagus |
|  |  | Gastritis and duodenitis |
|  |  | Gastro-oesophageal reflux disease |
|  |  | Oesophagitis and oesophageal ulcer |
|  | Inflammatory bowel disease | Crohn's disease |
|  |  | Ulcerative colitis |
|  | Coeliac disease |  |
|  | Diverticular disease of intestine (acute and chronic) |  |
|  | Fatty liver |  |
|  | Irritable bowel syndrome |  |
|  | Peptic ulcer disease |  |
|  | Hearing loss |  |
|  | Meniere disease |  |
|  | Addison’s disease |  |
|  | Cystic fibrosis |  |
|  | Hypo or hyperthyroidism |  |
|  | Type 1 diabetes |  |
|  | Type 2 diabetes |  |
|  | Diabetes not otherwise specified |  |
|  | Glaucoma |  |
|  | Macular degeneration |  |
|  | Visual impairment and blindness |  |
|  | Chronic renal disease | Chronic kidney disease |
|  |  | End stage renal disease |
|  |  | Glomerulonephritis |
|  |  | Tubulo-interstitial nephritis |
|  | Erectile dysfunction |  |
|  | Hyperplasia of prostate |  |
|  | Non-acute cystitis |  |
|  | Urinary incontinence |  |
|  | Allergic and chronic rhinitis |  |
|  | Asbestosis |  |
|  | Asthma |  |
|  | Bronchiectasis |  |
|  | Chronic obstructive pulmonary disease |  |
|  | Sleep apnoea |  |
|  | Iron and vitamin deficiency anaemia | Folate deficiency anaemia |
|  |  | Iron deficiency anaemia |
|  |  | Vitamin B12 deficiency anaemia |
|  | Immunodeficiencies |  |
|  | Sarcoidosis |  |
|  | Sickle-cell anaemia |  |
|  | Thalassaemia |  |
|  | HIV |  |
|  | Tuberculosis |  |
| Physical | Inflammatory arthritis and other inflammatory conditions | Ankylosing spondylitis |
|  |  | Juvenile arthritis |
|  |  | Lupus erythematosus (local and systemic) |
|  |  | Polymyalgia rheumatica |
|  |  | Postinfective and reactive arthropathies |
|  |  | Psoriatic arthropathy |
|  |  | Rheumatoid arthritis |
|  |  | Systemic sclerosis |
|  | Osteoporosis and vertebral crush fractures | Collapsed vertebra |
|  |  | Osteoporosis |
|  | Gout |  |
|  | Osteoarthritis (excl spine) |  |
|  | Spinal stenosis |  |
|  | Peripheral or autonomic neuropathy | Diabetic neurological complications |
|  |  | Disorders of autonomic nervous system |
|  |  | Peripheral neuropathies (excluding cranial nerve and carpal tunnel syndromes) |
|  | Cerebral palsy |  |
|  | Epilepsy |  |
|  | Migraine |  |
|  | Motor neurone disease |  |
|  | Multiple sclerosis |  |
|  | Myasthenia gravis |  |
|  | Parkinson's disease |  |
|  | Post-viral fatigue syndrome, neurasthenia and fibromyalgia |  |
|  | Down's syndrome |  |
|  | Psoriasis |  |
|  | Dementia |  |
| Mental | Alcohol problems |  |
|  | Anorexia and bulimia nervosa |  |
|  | Anxiety disorders |  |
|  | Autism and Asperger's syndrome |  |
|  | Bipolar affective disorder and mania |  |
|  | Intellectual disability |  |
|  | Obsessive-compulsive disorder |  |
|  | Other psychoactive substance misuse |  |
|  | Post-traumatic stress disorder |  |
|  | Schizophrenia, schizotypal and delusional disorders |  |

Abbreviation: CNS, central nervous system

### Table S3: Hazard ratio for the association of baseline depression, age, sex, sociodemographic, baseline comorbidities, and lifestyle factors with incident CHD (n=157,028)

| **Variables** |  | **Hazard Ratio (95% CI)** | | | |
| --- | --- | --- | --- | --- | --- |
|  |  | **Unadjusted** | **Model 1*** | **Model 2**** | **Model 3***** |
| **Depression** |  | 1.15 (1.08, 1.22) | 1.36 (1.28, 1.44) | 1.33 (1.25, 1.41) | 1.00 (0.94, 1.07) |
| **Age at baseline (years) ^a^** |  |  | 1.08 (1.07, 1.08) | 1.08 (1.07, 1.08) | 1.06 (1.06, 1.07) |
| **Age^2 at baseline ^b^** |  |  | 0.999 (0.999, 0.999) | 0.999 (0.998, 0.999) | 0.999 (0.998, 0.999) |
| **Sex (Ref: Female)** | Male |  | 2.23 (2.12, 2.34) | 2.22 (2.11, 2.33) | 2.25 (2.14, 2.36) |
| **Ethnicity (Ref: White)** | South Asian |  |  | 1.60 (1.34, 1.91) | 1.40 (1.17, 1.67) |
|  | Ethnic Minority Groups |  |  | 1.05 (0.89, 1.24) | 0.98 (0.82, 1.16) |
| **Country (Ref: England)** | Scotland |  |  | 0.75 (0.70, 0.81) | 0.78 (0.72, 0.83) |
|  | Wales |  |  | 0.77 (0.71, 0.84) | 0.73 (0.68, 0.79) |
| **Townsend Deprivation Index (Ref: 1)** | 2 |  |  | 1.10 (0.99, 1.23) | 1.09 (0.98, 1.21) |
|  | 3 |  |  | 1.06 (0.95, 1.18) | 1.03 (0.93, 1.15) |
|  | 4 |  |  | 1.15 (1.03, 1.28) | 1.12 (1.00, 1.24) |
|  | 5 |  |  | 1.15 (1.03, 1.27) | 1.09 (0.98, 1.21) |
|  | 6 |  |  | 1.13 (1.02, 1.26) | 1.06 (0.95, 1.18) |
|  | 7 |  |  | 1.17 (1.05, 1.30) | 1.07 (0.96, 1.19) |
|  | 8 |  |  | 1.28 (1.15, 1.42) | 1.12 (1.01, 1.25) |
|  | 9 |  |  | 1.35 (1.21, 1.50) | 1.12 (1.01, 1.25) |
|  | 10 (most deprived) |  |  | 1.69 (1.52, 1.88) | 1.24 (1.11, 1.38) |
| **No. of baseline conditions ^c^** |  |  |  |  | 1.18 (1.16, 1.20) |
| **No. of baseline conditions^2 ^d^** |  |  |  |  | 0.995 (0.991, 0.998) |
| **Smoking (Ref: Never)** | Previous |  |  |  | 1.09 (1.03, 1.15) |
|  | Current |  |  |  | 1.55 (1.44, 1.67) |
| **Alcohol intake (Ref: Daily or almost daily)** | Never |  |  |  | 1.42 (1.29, 1.56) |
|  | Special occasions only |  |  |  | 1.33 (1.22, 1.45) |
|  | 1-3 times a month |  |  |  | 1.23 (1.13, 1.35) |
|  | 1-2 times a week |  |  |  | 1.18 (1.10, 1.27) |
|  | 3 or 4 times a week |  |  |  | 1.03 (0.96, 1.10) |
| **Low physical activity (Ref: No)** | Yes |  |  |  | 1.25 (1.17, 1.34) |
| **Sleep disturbance (Ref: Never/ Rarely)** | Sometimes |  |  |  | 1.05 (0.99, 1.11) |
|  | Usually |  |  |  | 1.11 (1.04, 1.18) |
| **BMI (kg/m^2^) (Ref: <25)** | 25-29.9 |  |  |  | 1.31 (1.23, 1.39) |
|  | 30-34.9 |  |  |  | 1.51 (1.41, 1.62) |
|  | ≥35 |  |  |  | 1.61 (1.46, 1.76) |

Abbreviation: BMI, Body Mass Index

^a^ Scaled variable, Age fitted as (Age - mean Age)

^b^ Quadratic term for Age

^c^ Scaled variable, No. of baseline conditions fitted as (No. of baseline Conditions – mean No. of baseline Conditions)

^d^ Quadratic term for the No. of baseline conditions

* Adjusted for age and sex

** Further adjusted for ethnicity, country, and Townsend Deprivation Index

*** Further adjusted for no. of baseline conditions, smoking, alcohol intake, low physical activity, insomnia, and BMI

### Table S4: Hazard ratio for the association of baseline depression, age, sex, sociodemographic, baseline comorbidities, and lifestyle factors with incident PAD (n=167,125)

| **Variables** |  | **Hazard Ratio (95% CI)** | | | |
| --- | --- | --- | --- | --- | --- |
|  |  | **Unadjusted** | **Model 1*** | **Model 2**** | **Model 3***** |
| **Depression** |  | 1.32 (1.15, 1.52) | 1.58 (1.37, 1.83) | 1.48 (1.28, 1.70) | 0.89 (0.77, 1.04) |
| **Age at baseline (years) ^a^** |  |  | 1.11 (1.09, 1.12) | 1.11 (1.10, 1.12) | 1.09 (1.08, 1.10) |
| **Age^2 at baseline ^b^** |  |  | 0.999 (0.997, 1.000) | 0.998 (0.997, 1.000) | 0.998 (0.997, 1.000) |
| **Sex (Ref: Female)** | Male |  | 2.38 (2.10, 2.70) | 2.37 (2.09, 2.68) | 2.07 (1.81, 2.35) |
| **Ethnicity (Ref: White)** | South Asian |  |  | 1.56 (1.04, 2.35) | 1.56 (1.03, 2.37) |
|  | Ethnic Minority Groups |  |  | 0.91 (0.59, 1.40) | 0.87 (0.56, 1.35) |
| **Country (Ref: England)** | Scotland |  |  | 0.90 (0.76, 1.07) | 0.91 (0.77, 1.08) |
|  | Wales |  |  | 0.97 (0.80, 1.17) | 0.88 (0.72, 1.06) |
| **Townsend Deprivation Index (Ref: 1)** | 2 |  |  | 0.86 (0.63, 1.18) | 0.84 (0.61, 1.15) |
|  | 3 |  |  | 1.05 (0.78, 1.43) | 1.00 (0.74, 1.35) |
|  | 4 |  |  | 1.09 (0.80, 1.47) | 1.02 (0.75, 1.38) |
|  | 5 |  |  | 1.28 (0.96, 1.71) | 1.14 (0.85, 1.52) |
|  | 6 |  |  | 1.24 (0.93, 1.67) | 1.05 (0.79, 1.42) |
|  | 7 |  |  | 1.51 (1.13, 2.01) | 1.15 (0.86, 1.53) |
|  | 8 |  |  | 1.94 (1.48, 2.56) | 1.36 (1.03, 1.79) |
|  | 9 |  |  | 2.19 (1.67, 2.88) | 1.32 (1.00, 1.74) |
|  | 10 (most deprived) |  |  | 3.31 (2.55, 4.30) | 1.54 (1.18, 2.02) |
| **No. of baseline conditions ^c^** |  |  |  |  | 1.29 (1.24, 1.34) |
| **No. of baseline conditions^2 ^d^** |  |  |  |  | 0.990 (0.984, 0.996) |
| **Smoking (Ref: Never)** | Previous |  |  |  | 1.86 (1.59, 2.17) |
|  | Current |  |  |  | 6.88 (5.85, 8.08) |
| **Alcohol intake (Ref: Daily or almost daily)** | Never |  |  |  | 1.32 (1.06, 1.63) |
|  | Special occasions only |  |  |  | 1.10 (0.89, 1.36) |
|  | 1-3 times a month |  |  |  | 0.96 (0.76, 1.21) |
|  | 1-2 times a week |  |  |  | 0.96 (0.81, 1.14) |
|  | 3 or 4 times a week |  |  |  | 0.89 (0.74, 1.06) |
| **Low physical activity (Ref: No)** | Yes |  |  |  | 1.52 (1.31, 1.76) |
| **Sleep disturbance (Ref: Never/ Rarely)** | Sometimes |  |  |  | 1.00 (0.85, 1.17) |
|  | Usually |  |  |  | 1.07 (0.91, 1.26) |
| **BMI (kg/m^2^) (Ref: <25)** | 25-29.9 |  |  |  | 0.87 (0.75, 1.01) |
|  | 30-34.9 |  |  |  | 0.97 (0.82, 1.15) |
|  | ≥35 |  |  |  | 1.07 (0.86, 1.34) |

Abbreviation: BMI, Body Mass Index

^a^ Scaled variable, Age fitted as (Age - mean Age)

^b^ Quadratic term for Age

^c^ Scaled variable, No. of baseline conditions fitted as (No. of baseline Conditions – mean No. of baseline Conditions)

^d^ Quadratic term for the No. of baseline conditions

* Adjusted for age and sex

** Further adjusted for ethnicity, country, and Townsend Deprivation Index

*** Further adjusted for no. of baseline conditions, smoking, alcohol intake, low physical activity, insomnia, and BMI

### Table S5: Hazard ratio for the association of baseline depression, age, sex, sociodemographic, baseline comorbidities, and lifestyle factors with incident T2D (n=161,095)

| **Variables** |  | **Hazard Ratio (95% CI)** | | | |
| --- | --- | --- | --- | --- | --- |
|  |  | **Unadjusted** | **Model 1*** | **Model 2**** | **Model 3***** |
| **Depression** |  | 1.27 (1.19, 1.35) | 1.42 (1.34, 1.52) | 1.37 (1.29, 1.46) | 1.01 (0.94, 1.08) |
| **Age at baseline (years) ^a^** |  |  | 1.04 (1.03, 1.04) | 1.05 (1.04, 1.05) | 1.03 (1.03, 1.04) |
| **Age^2 at baseline ^b^** |  |  | 0.999 (0.999, 1.000) | 0.999 (0.998, 0.999) | 0.999 (0.999, 1.000) |
| **Sex (Ref: Female)** | Male |  | 1.83 (1.74, 1.93) | 1.82 (1.72, 1.91) | 1.95 (1.84, 2.06) |
| **Ethnicity (Ref: White)** | South Asian |  |  | 3.56 (3.11, 4.06) | 3.23 (2.82, 3.71) |
|  | Ethnic Minority Groups |  |  | 2.49 (2.21, 2.82) | 2.37 (2.09, 2.68) |
| **Country (Ref: England)** | Scotland |  |  | 0.82 (0.76, 0.89) | 0.83 (0.77, 0.90) |
|  | Wales |  |  | 1.12 (1.04, 1.21) | 0.99 (0.91, 1.07) |
| **Townsend Deprivation Index (Ref: 1)** | 2 |  |  | 0.95 (0.84, 1.09) | 0.93 (0.82, 1.06) |
|  | 3 |  |  | 1.05 (0.93, 1.20) | 0.98 (0.87, 1.12) |
|  | 4 |  |  | 1.15 (1.01, 1.31) | 1.07 (0.94, 1.21) |
|  | 5 |  |  | 1.22 (1.08, 1.38) | 1.07 (0.94, 1.21) |
|  | 6 |  |  | 1.30 (1.15, 1.47) | 1.10 (0.98, 1.25) |
|  | 7 |  |  | 1.34 (1.19, 1.52) | 1.08 (0.96, 1.23) |
|  | 8 |  |  | 1.56 (1.38, 1.75) | 1.18 (1.05, 1.34) |
|  | 9 |  |  | 1.82 (1.61, 2.04) | 1.25 (1.11, 1.41) |
|  | 10 (most deprived) |  |  | 2.34 (2.08, 2.63) | 1.41 (1.25, 1.59) |
| **No. of baseline conditions ^c^** |  |  |  |  | 1.13 (1.11, 1.15) |
| **No. of baseline conditions^2 ^d^** |  |  |  |  | 0.991 (0.988, 0.994) |
| **Smoking (Ref: Never)** | Previous |  |  |  | 1.10 (1.04, 1.17) |
|  | Current |  |  |  | 1.53 (1.41, 1.65) |
| **Alcohol intake (Ref: Daily or almost daily)** | Never |  |  |  | 1.67 (1.51, 1.85) |
|  | Special occasions only |  |  |  | 1.62 (1.48, 1.78) |
|  | 1-3 times a month |  |  |  | 1.38 (1.25, 1.53) |
|  | 1-2 times a week |  |  |  | 1.17 (1.07, 1.27) |
|  | 3 or 4 times a week |  |  |  | 1.07 (0.98, 1.17) |
| **Low physical activity (Ref: No)** | Yes |  |  |  | 1.25 (1.17, 1.34) |
| **Sleep disturbance (Ref: Never/ Rarely)** | Sometimes |  |  |  | 1.12 (1.04, 1.20) |
|  | Usually |  |  |  | 1.17 (1.09, 1.26) |
| **BMI (kg/m^2^) (Ref: <25)** | 25-29.9 |  |  |  | 2.97 (2.68, 3.29) |
|  | 30-34.9 |  |  |  | 7.04 (6.35, 7.80) |
|  | ≥35 |  |  |  | 13.87 (12.44, 15.46) |

Abbreviation: BMI, Body Mass Index

^a^ Scaled variable, Age fitted as (Age - mean Age)

^b^ Quadratic term for Age

^c^ Scaled variable, No. of baseline conditions fitted as (No. of baseline Conditions – mean No. of baseline Conditions)

^d^ Quadratic term for the No. of baseline conditions

* Adjusted for age and sex

** Further adjusted for ethnicity, country, and Townsend Deprivation Index

*** Further adjusted for no. of baseline conditions, smoking, alcohol intake, low physical activity, insomnia, and BMI

### Table S6: Hazard ratio for the association of baseline depression, age, sex, sociodemographic, baseline comorbidities, and lifestyle factors with IBD (n=166,045)

| **Variables** |  | **Hazard Ratio (95% CI)** | | | |
| --- | --- | --- | --- | --- | --- |
|  |  | **Unadjusted** | **Model 1*** | **Model 2**** | **Model 3***** |
| **Depression** |  | 1.30 (1.07, 1.58) | 1.34 (1.10, 1.63) | 1.30 (1.06, 1.58) | 1.00 (0.81, 1.23) |
| **Age at baseline (years) ^a^** |  |  | 1.01 (1.00, 1.02) | 1.02 (1.01, 1.03) | 1.00 (0.99, 1.01) |
| **Age^2 at baseline ^b^** |  |  | 1.000 (0.999, 1.002) | 1.000 (0.999, 1.002) | 1.000 (0.999, 1.001) |
| **Sex (Ref: Female)** | Male |  | 1.18 (1.00, 1.38) | 1.17 (0.99, 1.37) | 1.12 (0.95, 1.33) |
| **Ethnicity (Ref: White)** | South Asian |  |  | 1.79 (1.08, 2.96) | 1.79 (1.06, 3.00) |
|  | Ethnic Minority Groups |  |  | 1.00 (0.59, 1.68) | 0.99 (0.59, 1.67) |
| **Country (Ref: England)** | Scotland |  |  | 0.95 (0.75, 1.20) | 1.00 (0.79, 1.26) |
|  | Wales |  |  | 1.08 (0.85, 1.38) | 1.06 (0.83, 1.35) |
| **Townsend Deprivation Index (Ref: 1)** | 2 |  |  | 1.41 (0.95, 2.11) | 1.40 (0.94, 2.10) |
|  | 3 |  |  | 1.54 (1.04, 2.29) | 1.52 (1.02, 2.25) |
|  | 4 |  |  | 1.37 (0.91, 2.07) | 1.34 (0.89, 2.02) |
|  | 5 |  |  | 1.29 (0.86, 1.94) | 1.25 (0.83, 1.88) |
|  | 6 |  |  | 1.52 (1.02, 2.26) | 1.45 (0.97, 2.15) |
|  | 7 |  |  | 1.61 (1.08, 2.39) | 1.48 (1.00, 2.21) |
|  | 8 |  |  | 1.74 (1.18, 2.58) | 1.57 (1.06, 2.32) |
|  | 9 |  |  | 1.79 (1.21, 2.65) | 1.53 (1.03, 2.27) |
|  | 10 (most deprived) |  |  | 2.44 (1.67, 3.58) | 1.92 (1.30, 2.84) |
| **No. of baseline conditions ^c^** |  |  |  |  | 1.16 (1.10, 1.22) |
| **No. of baseline conditions^2 ^d^** |  |  |  |  | 0.998 (0.990, 1.007) |
| **Smoking (Ref: Never)** | Previous |  |  |  | 1.53 (1.28, 1.82) |
|  | Current |  |  |  | 1.67 (1.30, 2.15) |
| **Alcohol intake (Ref: Daily or almost daily)** | Never |  |  |  | 1.14 (0.82, 1.59) |
|  | Special occasions only |  |  |  | 1.34 (1.01, 1.78) |
|  | 1-3 times a month |  |  |  | 0.82 (0.59, 1.14) |
|  | 1-2 times a week |  |  |  | 1.04 (0.82, 1.32) |
|  | 3 or 4 times a week |  |  |  | 1.01 (0.79, 1.30) |
| **Low physical activity (Ref: No)** | Yes |  |  |  | 0.90 (0.70, 1.16) |
| **Sleep disturbance (Ref: Never/ Rarely)** | Sometimes |  |  |  | 1.03 (0.84, 1.28) |
|  | Usually |  |  |  | 1.12 (0.89, 1.41) |
| **BMI (kg/m^2^) (Ref: <25)** | 25-29.9 |  |  |  | 0.97 (0.80, 1.18) |
|  | 30-34.9 |  |  |  | 0.98 (0.77, 1.24) |
|  | ≥35 |  |  |  | 0.87 (0.63, 1.21) |

Abbreviation: BMI, Body Mass Index

^a^ Scaled variable, Age fitted as (Age - mean Age)

^b^ Quadratic term for Age

^c^ Scaled variable, No. of baseline conditions fitted as (No. of baseline Conditions – mean No. of baseline Conditions)

^d^ Quadratic term for the No. of baseline conditions

* Adjusted for age and sex

** Further adjusted for ethnicity, country, and Townsend Deprivation Index

*** Further adjusted for no. of baseline conditions, smoking, alcohol intake, low physical activity, insomnia, and BMI

### Table S7: Hazard ratio for the association of baseline depression, age, sex, sociodemographic, baseline comorbidities, and lifestyle factors with incident PD (n=168,347)

| **Variables** |  | **Hazard Ratio (95% CI)** | | | |
| --- | --- | --- | --- | --- | --- |
|  |  | **Unadjusted** | **Model 1*** | **Model 2**** | **Model 3***** |
| **Depression** |  | 1.29 (1.05, 1.57) | 1.52 (1.25, 1.86) | 1.53 (1.25, 1.87) | 1.45 (1.18, 1.79) |
| **Age at baseline (years) ^a^** |  |  | 1.14 (1.12, 1.16) | 1.14 (1.12, 1.16) | 1.14 (1.12, 1.16) |
| **Age^2 at baseline ^b^** |  |  | 0.998 (0.996, 1.000) | 0.998 (0.996, 1.000) | 0.998 (0.996, 1.000) |
| **Sex (Ref: Female)** | Male |  | 2.00 (1.69, 2.37) | 2.00 (1.69, 2.37) | 2.08 (1.74, 2.49) |
| **Ethnicity (Ref: White)** | South Asian |  |  | 0.82 (0.34, 1.98) | 0.60 (0.25, 1.47) |
|  | Ethnic Minority Groups |  |  | 1.54 (0.86, 2.73) | 1.35 (0.76, 2.41) |
| **Country (Ref: England)** | Scotland |  |  | 0.92 (0.72, 1.17) | 0.93 (0.73, 1.18) |
|  | Wales |  |  | 0.94 (0.72, 1.21) | 0.94 (0.72, 1.21) |
| **Townsend Deprivation Index (Ref: 1)** | 2 |  |  | 0.74 (0.52, 1.04) | 0.74 (0.52, 1.04) |
|  | 3 |  |  | 0.92 (0.66, 1.29) | 0.92 (0.66, 1.28) |
|  | 4 |  |  | 0.95 (0.68, 1.33) | 0.95 (0.68, 1.33) |
|  | 5 |  |  | 0.85 (0.61, 1.19) | 0.85 (0.61, 1.19) |
|  | 6 |  |  | 0.61 (0.42, 0.89) | 0.61 (0.42, 0.89) |
|  | 7 |  |  | 0.72 (0.50, 1.04) | 0.73 (0.51, 1.05) |
|  | 8 |  |  | 0.83 (0.58, 1.19) | 0.84 (0.59, 1.20) |
|  | 9 |  |  | 1.10 (0.79, 1.54) | 1.12 (0.80, 1.57) |
|  | 10 (most deprived) |  |  | 0.65 (0.44, 0.98) | 0.67 (0.44, 1.00) |
| **No. of baseline conditions ^c^** |  |  |  |  | 1.03 (0.98, 1.09) |
| **No. of baseline conditions^2 ^d^** |  |  |  |  | 1.005 (0.997, 1.013) |
| **Smoking (Ref: Never)** | Previous |  |  |  | 0.82 (0.68, 0.97) |
|  | Current |  |  |  | 0.50 (0.34, 0.73) |
| **Alcohol intake (Ref: Daily or almost daily)** | Never |  |  |  | 1.51 (1.10, 2.06) |
|  | Special occasions only |  |  |  | 1.16 (0.85, 1.58) |
|  | 1-3 times a month |  |  |  | 1.23 (0.91, 1.67) |
|  | 1-2 times a week |  |  |  | 0.95 (0.74, 1.22) |
|  | 3 or 4 times a week |  |  |  | 0.97 (0.76, 1.25) |
| **Low physical activity (Ref: No)** | Yes |  |  |  | 1.07 (0.81, 1.40) |
| **Sleep disturbance (Ref: Never/ Rarely)** | Sometimes |  |  |  | 0.89 (0.73, 1.09) |
|  | Usually |  |  |  | 0.86 (0.68, 1.08) |
| **BMI (kg/m^2^) (Ref: <25)** | 25-29.9 |  |  |  | 1.21 (0.99, 1.48) |
|  | 30-34.9 |  |  |  | 0.99 (0.77, 1.28) |
|  | ≥35 |  |  |  | 0.82 (0.55, 1.22) |

Abbreviation: BMI, Body Mass Index

^a^ Scaled variable, Age fitted as (Age - mean Age)

^b^ Quadratic term for Age

^c^ Scaled variable, No. of baseline conditions fitted as (No. of baseline Conditions – mean No. of baseline Conditions)

^d^ Quadratic term for the No. of baseline conditions

* Adjusted for age and sex

** Further adjusted for ethnicity, country, and Townsend Deprivation Index

*** Further adjusted for no. of baseline conditions, smoking, alcohol intake, low physical activity, insomnia, and BMI

### Table S8: Hazard ratio for the association of baseline depression, age, sex, sociodemographic, baseline comorbidities, and lifestyle factors with incident IA (n=163,189)

| **Variables** |  | **Hazard Ratio (95% CI)** | | | |
| --- | --- | --- | --- | --- | --- |
|  |  | **Unadjusted** | **Model 1*** | **Model 2**** | **Model 3***** |
| **Depression** |  | 1.40 (1.28, 1.53) | 1.37 (1.25, 1.50) | 1.37 (1.25, 1.50) | 1.07 (0.97, 1.17) |
| **Age at baseline (years) ^a^** |  |  | 1.05 (1.05, 1.06) | 1.06 (1.05, 1.06) | 1.04 (1.03, 1.05) |
| **Age^2 at baseline ^b^** |  |  | 1.001 (1.000, 1.001) | 1.001 (1.000, 1.001) | 1.000 (1.000, 1.001) |
| **Sex (Ref: Female)** | Male |  | 0.69 (0.64, 0.75) | 0.69 (0.63, 0.74) | 0.66 (0.61, 0.72) |
| **Ethnicity (Ref: White)** | South Asian |  |  | 1.62 (1.22, 2.14) | 1.46 (1.10, 1.95) |
|  | Ethnic Minority Groups |  |  | 1.24 (0.97, 1.60) | 1.21 (0.94, 1.55) |
| **Country (Ref: England)** | Scotland |  |  | 0.77 (0.68, 0.86) | 0.80 (0.71, 0.90) |
|  | Wales |  |  | 0.88 (0.78, 0.99) | 0.84 (0.75, 0.95) |
| **Townsend Deprivation Index (Ref: 1)** | 2 |  |  | 0.89 (0.75, 1.04) | 0.88 (0.75, 1.04) |
|  | 3 |  |  | 0.92 (0.78, 1.08) | 0.90 (0.77, 1.06) |
|  | 4 |  |  | 0.98 (0.84, 1.16) | 0.96 (0.82, 1.13) |
|  | 5 |  |  | 0.90 (0.76, 1.06) | 0.87 (0.74, 1.02) |
|  | 6 |  |  | 0.87 (0.74, 1.03) | 0.83 (0.70, 0.98) |
|  | 7 |  |  | 0.96 (0.81, 1.13) | 0.88 (0.75, 1.04) |
|  | 8 |  |  | 0.96 (0.81, 1.14) | 0.86 (0.73, 1.02) |
|  | 9 |  |  | 1.00 (0.84, 1.18) | 0.85 (0.72, 1.00) |
|  | 10 (most deprived) |  |  | 1.01 (0.85, 1.20) | 0.79 (0.66, 0.94) |
| **No. of baseline conditions ^c^** |  |  |  |  | 1.18 (1.15, 1.21) |
| **No. of baseline conditions^2 ^d^** |  |  |  |  | 0.992 (0.987, 0.996) |
| **Smoking (Ref: Never)** | Previous |  |  |  | 1.12 (1.03, 1.21) |
|  | Current |  |  |  | 1.44 (1.27, 1.63) |
| **Alcohol intake (Ref: Daily or almost daily)** | Never |  |  |  | 1.11 (0.95, 1.29) |
|  | Special occasions only |  |  |  | 1.09 (0.95, 1.25) |
|  | 1-3 times a month |  |  |  | 0.95 (0.82, 1.10) |
|  | 1-2 times a week |  |  |  | 0.95 (0.85, 1.07) |
|  | 3 or 4 times a week |  |  |  | 0.99 (0.88, 1.12) |
| **Low physical activity (Ref: No)** | Yes |  |  |  | 1.12 (1.00, 1.25) |
| **Sleep disturbance (Ref: Never/ Rarely)** | Sometimes |  |  |  | 1.06 (0.96, 1.18) |
|  | Usually |  |  |  | 1.16 (1.04, 1.30) |
| **BMI (kg/m^2^) (Ref: <25)** | 25-29.9 |  |  |  | 1.14 (1.04, 1.26) |
|  | 30-34.9 |  |  |  | 1.20 (1.07, 1.34) |
|  | ≥35 |  |  |  | 1.33 (1.15, 1.54) |

Abbreviation: BMI, Body Mass Index

^a^ Scaled variable, Age fitted as (Age - mean Age)

^b^ Quadratic term for Age

^c^ Scaled variable, No. of baseline conditions fitted as (No. of baseline Conditions – mean No. of baseline Conditions)

^d^ Quadratic term for the No. of baseline conditions

* Adjusted for age and sex

** Further adjusted for ethnicity, country, and Townsend Deprivation Index

*** Further adjusted for no. of baseline conditions, smoking, alcohol intake, low physical activity, insomnia, and BMI
